# More efficient and inclusive time-to-event trials with covariate adjustment: a simulation study

**DOI:** 10.1101/2022.04.15.22273871

**Authors:** Raphaëlle Momal, Paul Trichelair, Michael G.B. Blum, Félix Balazard

## Abstract

Adjustment for prognostic covariates increases the statistical power of randomized trials. The factors influencing increase of power are well-known for trials with continuous outcomes. Here, we study which factors influence power and sample size requirements in time-to-event trials. We consider both parametric simulations and simulations derived from the TCGA cohort of hepatocellular carcinoma (HCC) patients to assess how sample size requirements are reduced with covariate adjustment. Simulations demonstrate that the benefit of covariate adjustment increases with the prognostic performance of the adjustment covariate (C-index) and with the cumulative incidence of the event in the trial. For a covariate that has an intermediate prognostic performance (C-index=0.65), the reduction of sample size varies from 1.7% when cumulative incidence is of 10% to 26.5% when cumulative incidence is of 90%. Broadening eligibility criteria usually reduces statistical power while our simulations show that it can be maintained with adequate covariate adjustment. In a simulation of HCC trials, we find that the number of patients screened for eligibility can be divided by 2.7 when broadening eligibility criteria. Last, we find that the Cox-Snell 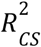 is a good approximation of the reduction in sample size requirements provided by covariate adjustment. This metric can be used in the design of time-to-event trials to determine sample size. Overall, more systematic adjustment for prognostic covariates leads to more efficient and inclusive clinical trials especially when cumulative incidence is large as in metastatic and advanced cancers.

**Key messages:**

- Covariate adjustment is a statistical technique that leverages prognostic scores within the statistical analysis of a trial. We study its benefits for time-to-event trials.
- Power gain achieved with covariate adjustment is determined by the prognostic performance of the covariate and by the cumulative incidence of events at the end of the follow-up period.
- Trials in indications with large cumulative incidence such as metastatic cancers can benefit from covariate adjustment to improve their statistical power.
- Covariate adjustment maintains statistical power in trials when eligibility criteria are broadened.

## Introduction

Adjustment for prognostic covariates improves precision and increases statistical power for treatment effect estimation in randomized clinical trials[1–4]. Randomization guarantees the validity of statistical analysis of randomized trials whether they are adjusted or unadjusted[5]. However, unadjusted analysis can be imprecise because of a large variability between patient outcomes that is explained by several baseline covariates. Covariate adjustment accounts for outcome variation between patients, leading to a more precise estimation of the treatment effect[1]. While adjustment on important covariates can correct for chance imbalance in important baseline covariates, adjustment covariates should be selected and prespecified at the trial design stage based on their prognostic value and not on any imbalance criterion assessed after randomization[6]. This methodological consensus is currently being translated into regulatory guidance: EMA published a guideline in 2015 and the FDA has issued a draft guidance in 2021[7,8]. Increase of precision when using covariate adjustment translates to a reduced sample size for reaching a target of statistical power, typically 80% in clinical trials.

For time-to-event trials that are frequent in oncology, we investigate to what extent trial and indication characteristics determine the impact of covariate adjustment on statistical power and on sample size requirements. These characteristics include the cumulative incidence of the event of interest at the end of follow-up, the prognostic performance of covariates and the censoring rate. Understanding the relationship between cumulative incidence and reduction in sample size helps prioritize the disease indications where covariate adjustment is the most impactful.

We also evaluate whether covariate adjustment can help to broaden trial eligibility criteria. Eligibility criteria in clinical trials can be too restrictive which leads to limited generalizability as well as difficulty in enrollment[10,11]. Beyond ensuring patient safety, restrictive eligibility might be used to ensure homogeneity in the trial population[12,13]. In non-small cell lung cancer, it was shown using observational cohorts that many inclusion criteria are superfluous as they restrict the potential enrollment of trials even though the treatment is as efficacious for the excluded patients as for the included patients[14]. As covariate adjustment allows to analytically compensate for the heterogeneity in the patient population, we investigate whether adequate covariate adjustment could allow to broaden eligibility criteria while maintaining statistical power.

To answer both of those questions, we use parametric simulations as well as semi-synthetic simulations based on data from patients with resected hepatocellular carcinoma (HCC). In parametric simulations, event times are simulated based on an extensive exploration of the parameter space. The semi-synthetic simulations are based on TCGA data[15]. The covariate of interest, which is used for adjustment, is a deep-learning model based on histological slides. This covariate named HCCnet captures a prognostic signal on overall survival for HCC after resection[16]. In both cases, the simulations rely on the proportional hazards assumption.

Last, we determine how sample size could be determined if the prognostic signal carried by the covariate is known a priori based on external data. For a continuous outcome, the Fleiss formula relates the sample size of the adjusted analysis (denoted *N*_*adj*_), which is required for a given statistical power, to the sample size of the unadjusted analysis (denoted *N*_0_). Denoting by *r*^2^ the proportion of variance of the outcome explained by the covariate, the Fleiss formula states that the sample size needed for the adjusted analyses is reduced by *r*^2^ compared to the unadjusted one *N*_*adj*_ = *N*_0_ (1 − *r*^2^) [9]. For instance a correlation *r* of 0.5 between a baseline covariate and the outcome translates to sample size requirements for the adjusted analysis reduced by 25% compared to the unadjusted analysis. For a time-to-event outcome, there is not a unique definition for the proportion of variation explained by a covariate. Different measures to compute the proportion of explained variance have been proposed for survival analysis [17,18]. Using the parametric simulations, we assess whether a measure extends the Fleiss formula for a time-to-event setting.

## Methods

### Parametric simulations based on a time-to-event model

Parametric simulations are performed to estimate the observed reduction of the sample size requirement and assess its relationship with a single adjustment covariate’s C-index and the cumulative incidence of the event. Other parameters of interest are the size of the treatment effect, the Weibull shape of the baseline hazard function and the drop-out rate. The simulations rely on the proportional hazard assumption.

Survival times are generated following the Weibull distribution with shape *w* and scale depending on the treatment hazard ratio *hr*, and on a standard Gaussian covariate *x*. Censored times T^*drop*^ are drawn from an exponential distribution with a specified drop-out rate *d*. Denoting *z* the treatment allocation variable, κ the intercept and θ the coefficient of *x*, this generative model can be formally summarized as follows for patient *i*:

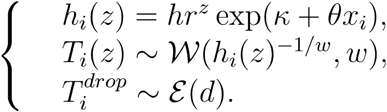

All patients remaining at risk at 5 years are censored at that time. The treatment allocation is independent of the covariate and there is the same number of patients in both arms. For each set of input parameters, the auxiliary parameters κ and θ are numerically optimized to reach a prespecified value Λ of the cumulative incidence in the control arm, and the C-index *C* evaluated on the whole trial population.

Once survival times are simulated, the presence of a treatment effect is tested in an unadjusted analysis and an analysis adjusted for the covariate using the Wald test for the treatment coefficient in a Cox regression. The statistical power for the unadjusted analysis and the adjusted analysis is estimated on a grid of sample sizes based on 10,000 numerical repetitions per sample size[19]. The resulting power curves give the sample sizes *N*_*adj*_ and *N*_*0*_ required to reach a power of 80% for both analyses, from which the reduction of sample size achieved with adjustment R^2^_obs_ is deduced (Figure 1). These simulations explore a wide range of parameter values (Table S1), allowing for an extensive study of R^2^_obs_ behavior as a function of the cumulative incidence Λ and c-index *C* in different settings of proportional hazards.

**Figure 1:**
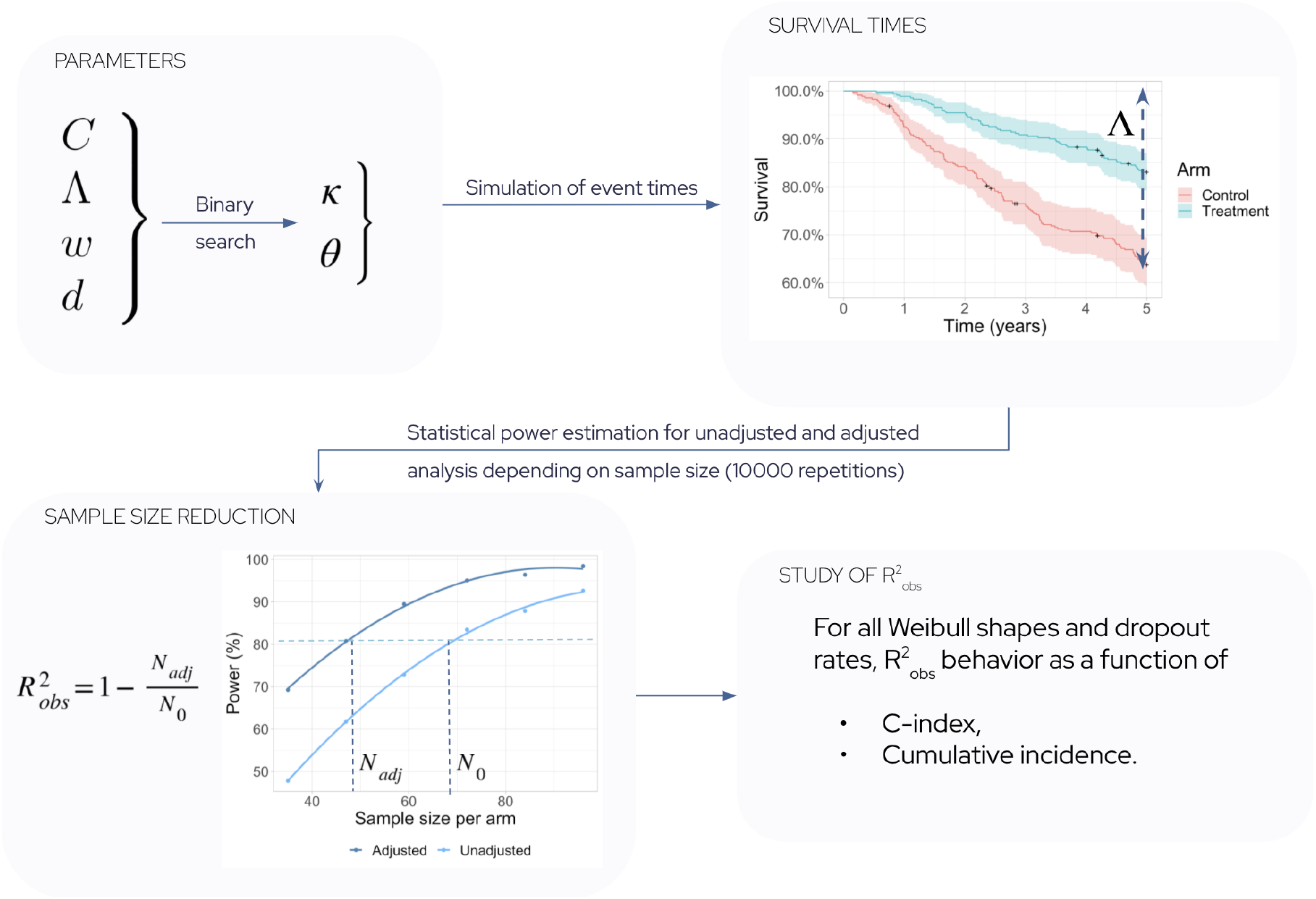
Workflow of parametric simulations. For a set of parameters corresponding to a clinical trial scenario,10,000 instances of clinical trials are simulated to estimate statistical power.

To indicate what are the most relevant indications for covariate adjustment, we provide estimates of the cumulative incidence Λ in the control arm for several oncology trials. Cumulative incidence is estimated by reading the value of the Kaplan-Meier curves published in the manuscript describing the trial results.

### Semi-synthetic simulations based on HCC data from TCGA

To consider simulations that mimic distributions of covariates found in clinical data, we also perform semi-synthetic simulations of resected HCC patients. The covariate used for adjustment is a prognostic score based on H&E images processed with the HCCnet deep learning algorithm[16]. The deep learning model was trained on another dataset than TCGA. We consider the prognostic scores of HCCnet applied on 328 patients with early stage HCC from the TCGA HCC dataset[15,16]. In the TCGA dataset, we have access to outcome measures including overall survival and 73 clinical variables with less than 50% of missing data in addition to the HCCnet prognostic covariate.

We impute all missing values among the 73 clinical variables having less than 50% of missing variables and more than one modality. For imputation, we use factorial analysis for mixed data (FAMD), a principal component method for data involving both continuous and categorical variables[20]. The imputed variables used as adjustment are tumor staging (1% missing values) and ECOG score which have 20% missing values. The imputed variables used as eligibility criteria in our simulation study are the ECOG score, the Child Pugh classification (33% missing), the macrovascular invasion (15% missing) and B or C hepatitis infection status (15% and 5% missing values respectively).

The simulations follow the same assumptions as the parametric model while preserving the observed survival curve and dependence of survival on covariates. To do so, a Cox model of overall survival is fitted on the available prognostic variables (tumor staging, ECOG score and the HCCnet variable). For each simulated patient, we sample her/his clinical covariates from TCGA. The hazard rate is defined as for parametric simulations except that there is a matrix *X* of covariates instead of a single covariate, and θ is replaced by 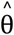 the vector of coefficients obtained from the fitted Cox model. The Weibull distribution is replaced by the empirical survival function that depends on the hazard rate and on the baseline survival function *Ŝ*_0_ estimated with Kaplan-Meier:

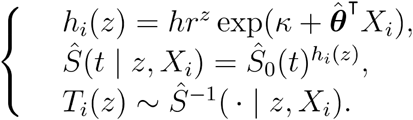

As before, κ is numerically optimized in order to set the incidence of outcome in the control arm to the observed incidence of outcome. Finally, all patients with events after 5 years are censored at that time.

We choose a sample size of 760 individuals as it is the average sample size of 4 ongoing trials for adjuvant treatment in early stage HCC[21–24]. The treatment effect size *hr* is set so that the estimated statistical power with adjustment for the clinical variables (tumor staging and ECOG score) is 80% for a sample size of 760 individuals. Randomization of the treatment assignment is stratified on tumor staging. To estimate the reduction of sample size obtained when adding HCCNet as adjustment covariate, we consider varying values of the sample size, find the minimal values where power reaches 80% and compute the relative reduction of sample size compared to the sample size of 760 individuals. Statistical power is estimated based on 5,000 repetitions.

### Effect of covariate adjustment when broadening eligibility criteria

Using the parametric simulations and the semi-synthetic simulations, we evaluate if the effect of covariate adjustment is changed when considering less restrictive inclusion criteria. For parametric simulations, the restricted inclusion criteria is based on the values of the prognostic covariate *X*. Only patients with values of *X* below 0, i.e. patients at lower risk, are included in the simulated trial with the most restrictive eligibility criteria. There are then 4 scenarios when combining the two possible eligibility criteria (all patients or restricted inclusion) and the two choices of adjustments (no adjustment or adjustment for *X*). Parameters of the simulations include the hazard ratio of the covariate, the intercept of the Cox model, the Weibull shape and the treatment hazard ratio which are set to θ = 3, κ = 0 *w* = 1, and *hr* = 0. 7 respectively. The observed cumulative incidence is 96.5% when all patients are included, and 93.5% for the restricted inclusion.

In the case of the HCC semi-synthetic simulations, we consider that including all TCGA patients selected for HCCnet validation is the less restrictive inclusion criteria and we define two additional levels of restricted eligibility criteria (Table 1). The mildly restrictive eligibility level has two inclusion criteria present in all 4 ongoing large trials for adjuvant treatment in early stage HCC[21–24]: only patients with a Child-Pugh score of A and with an ECOG status of 0 or 1 are included. The most restrictive eligibility criteria further restrict the ECOG status to 0 as in the STORM trial[25], exclude patients with a dual infection of hepatitis B and hepatitis C as in the KEYNOTE-937 trial[23] and exclude patients with macrovascular invasion as in the IMBRAVE050 trial[22]. We consider only the eligibility criteria that were available in the TCGA HCC dataset. There are therefore 6 different scenarios when combining the three levels of eligibility criteria and the two choices of adjustment: whether or not HCCnet is considered as an adjustment variable in addition to tumor staging and ECOG. In the scenario with the most restrictive eligibility levels, every patient has an ECOG of 0 and therefore the analyses are not adjusted for ECOG.

**Table 1:**
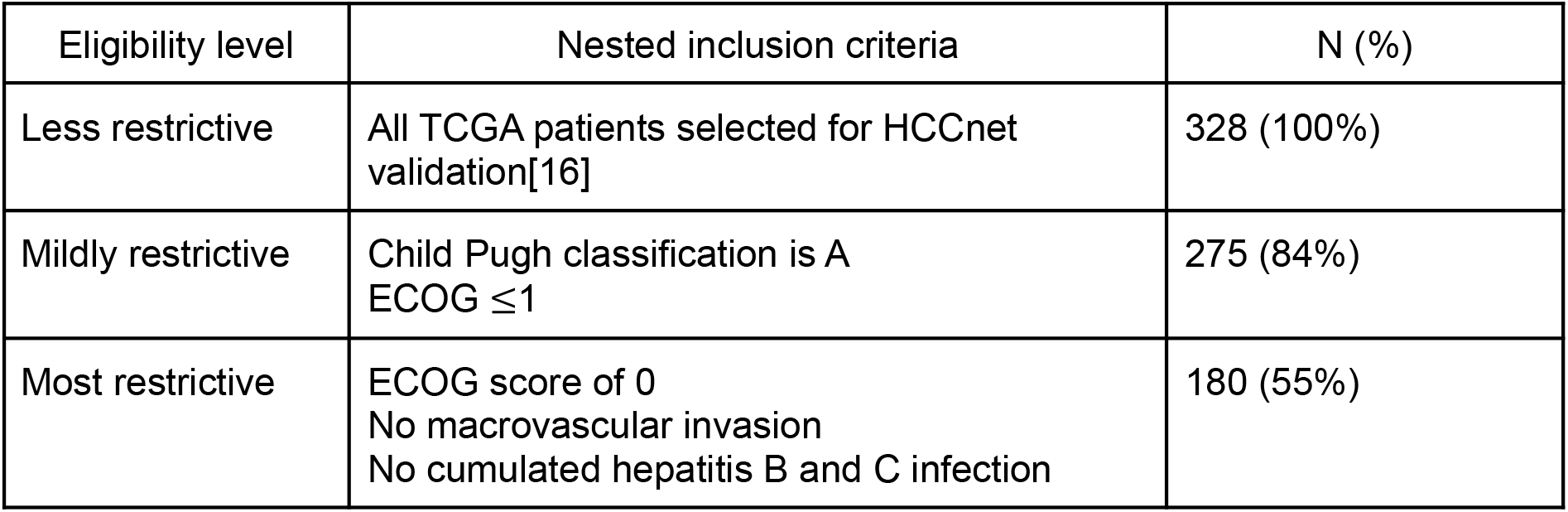
Definition of eligibility criteria used for the semi-synthetic simulations based on the TCGA dataset.

For both types of simulations, changing the inclusion criteria changes the number of events which affects statistical power directly. To provide a fair comparison between the methods with or without adjustment, we present the statistical power of the different scenarios as a function of the number of events. In both cases, no drop-out was added and 5000 repetitions were performed to evaluate statistical power. In both cases, patients at lower risk of the event are selected when we consider the more restrictive criteria. We also evaluate how broadening the eligibility criteria would impact the number of patients that need to be screened for enrollment to succeed.

### Proposed R^2^ measures for survival analysis

Several categories of measures have been proposed to extend the R^2^ measure to time-to-event data[17,18]. We consider explained variation (EV) and explained randomness (ER) measures. Explained variation measures are extensions of the proportion of explained variance that is used in linear regression. Explained randomness measures, on the other hand, are based on entropy measures and compare the quantity of information contained in models with and without the covariates of interest. In the simulations, we study the behavior of three EV measures: 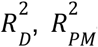 and 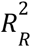 [26–28], and five ER measures: 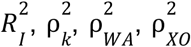 and 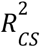 [26,27,29–31]. The proposed R^2^ measures are computed over the grid of simulation parameters and compared to the observed reduction in sample size.

## Results

### Evaluation of the factors impacting sample size reduction with parametric simulations

The parametric simulations show that the sample size reduction obtained with covariate adjustment varies between 0 and 80%. It increases as a function of the covariate prognostic performance measured with the C-index, and of cumulative incidence, which corresponds to the probability of an event (death, progression…) before the end of the follow-up period. When we consider a cumulative incidence of Λ = 10%, covariate adjustment reduces the sample size by 1.7% for a covariate with a C-index of 0.65, by 5.4% for a C-index of 0.75, and by 16.1% for a C-index is 0.85. For an intermediate value of Λ = 50%, the reduction is 11.7%, 35.4%, 64.9% for the three C-index values of 0.65, 0.75, and 0.85. For a high cumulative incidence value of Λ = 90%, the reduction is 26.5%, 54.1%, 80.5% for the same values of C-index (Figure 2).

**Figure 2:**
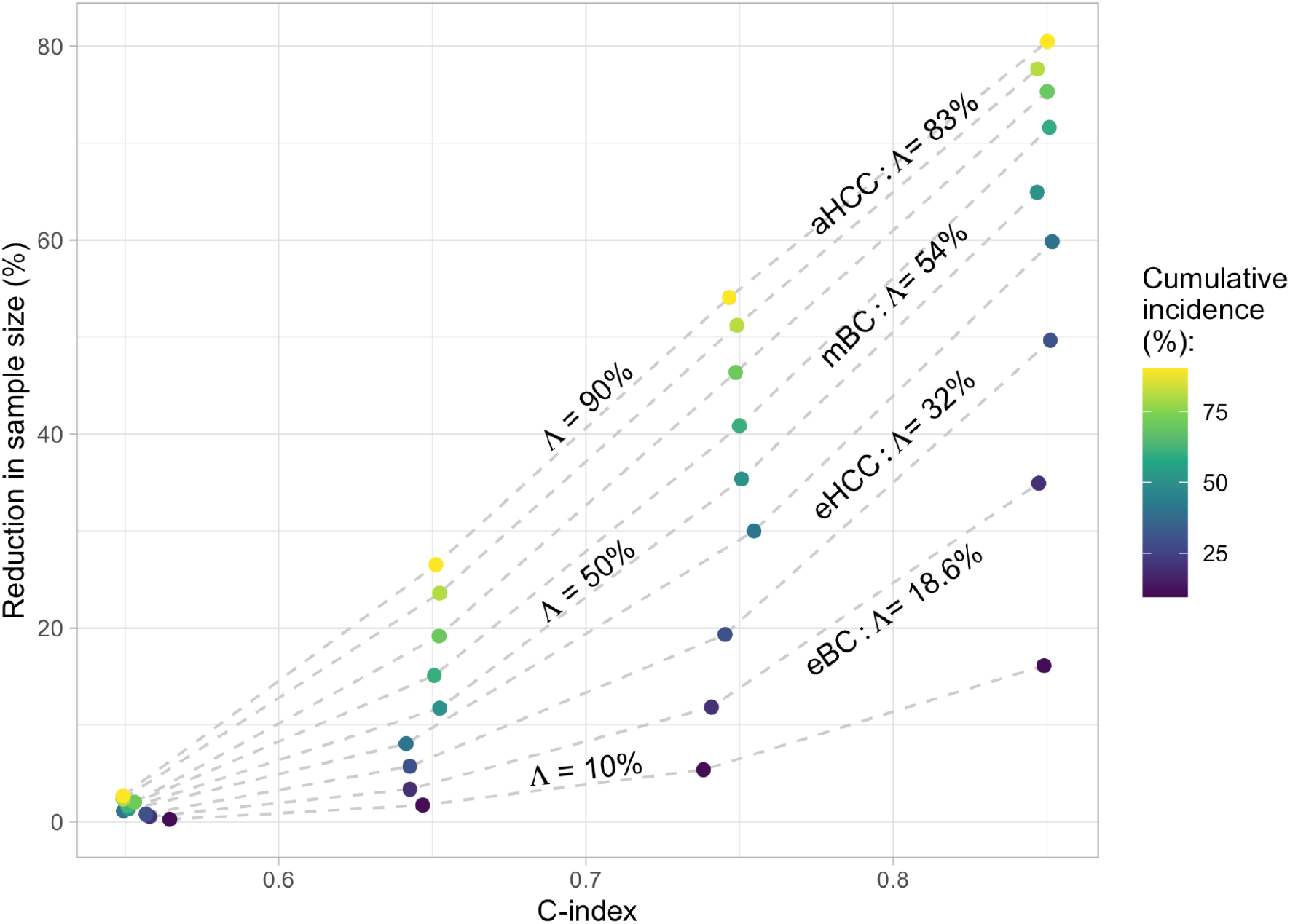
Reduction in sample size R^2^_obs_ as a function of the prognostic performance (C-index) of the covariate for a range of cumulative incidence values. Cumulative incidence Λ is measured at the end of the follow-up period. In the simulations, the hazard ratio is set at hr=0.7, the drop-out rate at d=0.01, and the shape parameter of the Weibull distribution at w=0.5. The cumulative incidence values that are provided for the breast cancer and HCC indications come from clinical trials selected in table 2. eBC: early breast cancer; eHCC: early resectable hepatocellular carcinoma; mBC: metastatic breast cancer; aHCC: advanced hepatocellular carcinoma.

Cumulative incidence values depend on indication, on the nature of the event (progression, death…) and on the duration of follow-up (Table 2). We find a wide range of values for cumulative incidence in several oncology trials. It ranges from 18.6% at 5 years for disease recurrence in early breast cancer to 98% at 3 years for death in metastatic pancreatic cancer (Table 2).

**Table 2:** Cumulative incidence of events of interest in the control arms of a selection of trials. For a given C-index of a prognostic covariate, the impact of covariate adjustment will be larger for indications with large cumulative incidence of events. HR+: Hormone receptor positive; PD-L1+: programmed death ligand 1 positive; NSCLC: non-small cell lung cancer

| Indication | Trial | Cumulative incidence $\Lambda$ in control arm |
| --- | --- | --- |
| HR+ early breast cancer (eBC) | BIG 1-98 [32]<br>Letrozole vs tamoxifen | Probability of disease recurrence at 5 years: 18.6% |
| HCC after resection of local ablation (eHCC) | STORM[25]<br>Sorafenib vs placebo | Probability of death at 5 years: 32% |
| Metastatic hormone-sensitive prostate cancer | ENZAMET[33]<br>Enzalutamide vs standard nonsteroidal antiandrogen therapy in addition to testosterone suppression | Probability of death at 4 years: 36% |
| PD-L1+ advanced NSCLC | KEYNOTE-024[34]<br>Pembrolizumab vs chemotherapy | Probability of death at 1.5 years: 50% |
| HR+ metastatic breast cancer in premenopausal patients (mBC) | MONALEESA-7[35]<br>Ribociclib vs placebo in addition to endocrine therapy | Probability of death at 3.5 years: 54% |
| Resected pancreatic cancer | PRODIGE 24[36]<br>Modified FOLFIRINOX vs gemcitabine | Probability of death at 5 years: 70% |
| Advanced HCC (aHCC) | CheckMate 459[37]<br>Nivolumab vs sorafenib | Probability of death at 3 years: 83% |
| Malignant pleural mesothelioma | CheckMate 743[38]<br>Nivolumab+Ipilimumab vs chemotherapy | Probability of death at 3 years: 85% |
| Metastatic pancreatic cancer | OXIPAN[39]<br>FOLFIRINOX vs gemcitabine | Probability of death at 3 years: 98% |

We find that other parameters of the simulations do not impact the reduction of sample size obtained with covariate adjustment. These additional parameters are the size of the treatment effect (hazard ratio), the Weibull shape parameter and the drop-out rate (Figure S1).

The drop-out rates of d=0.01 or d=0.1 result in different average censoring rates depending on the values taken by other parameters. The median censoring rate (computed over the set of other parameters’ value) before the end of follow-up was 3.7% when d=0.01 (min-max: 0.7%-4.8%) and 28.1% when d=0.1 (min-max: 5.4%-38.4%).

### Comparing semi-synthetic HCC simulations and parametric simulations

We consider semi-synthetic simulations based on the TGCA HCC cohort to evaluate power gain obtained with a deep learning variable. We find that adjusting on the deep learning covariate HCCnet, in addition to tumor staging and ECOG, reduces the required sample size by 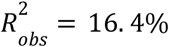 (figure S2). In terms of statistical power, the increase provided by the deep learning covariate is 6.4% for the sample size that provides a power of 80% when adjusting on ECOG and tumor staging only.

We evaluate the compatibility of this result with the results of the parametric simulations. The cumulative incidence of death in the HCC-TCGA population is 50% at 5 years. The Cox model with tumor staging and ECOG score has a C-index of 0.65 in the source population, while adding HCCnet results in a C-index of 0.72. We label by 1 the quantities associated with adjustment for the clinical variables (tumor staging and ECOG) and by 2 the quantities associated with the additional adjustment of the HCCnet covariate (tumor staging, ECOG and HCCnet). Applying Fleiss equation for the two adjustments using the 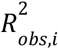 obtained with parametric simulations (Figure 2) we obtain

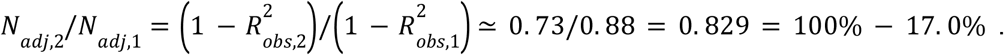

Therefore, results obtained with semi-synthetic simulations are coherent with the findings of the parametric simulations.

### Covariate adjustment when broadening eligibility criteria

When broadening eligibility criteria, statistical power of unadjusted analysis is reduced for a fixed number of events (Figure 3). By contrast, if we consider covariate adjustment in the parametric simulations, power is almost not affected by eligibility criteria. In the semi-synthetic simulations, we find that further adjusting on the additional HCCnet covariate reduces the extent to which power is affected by changes of eligibility criteria (Figure 3).

**Figure 3:**
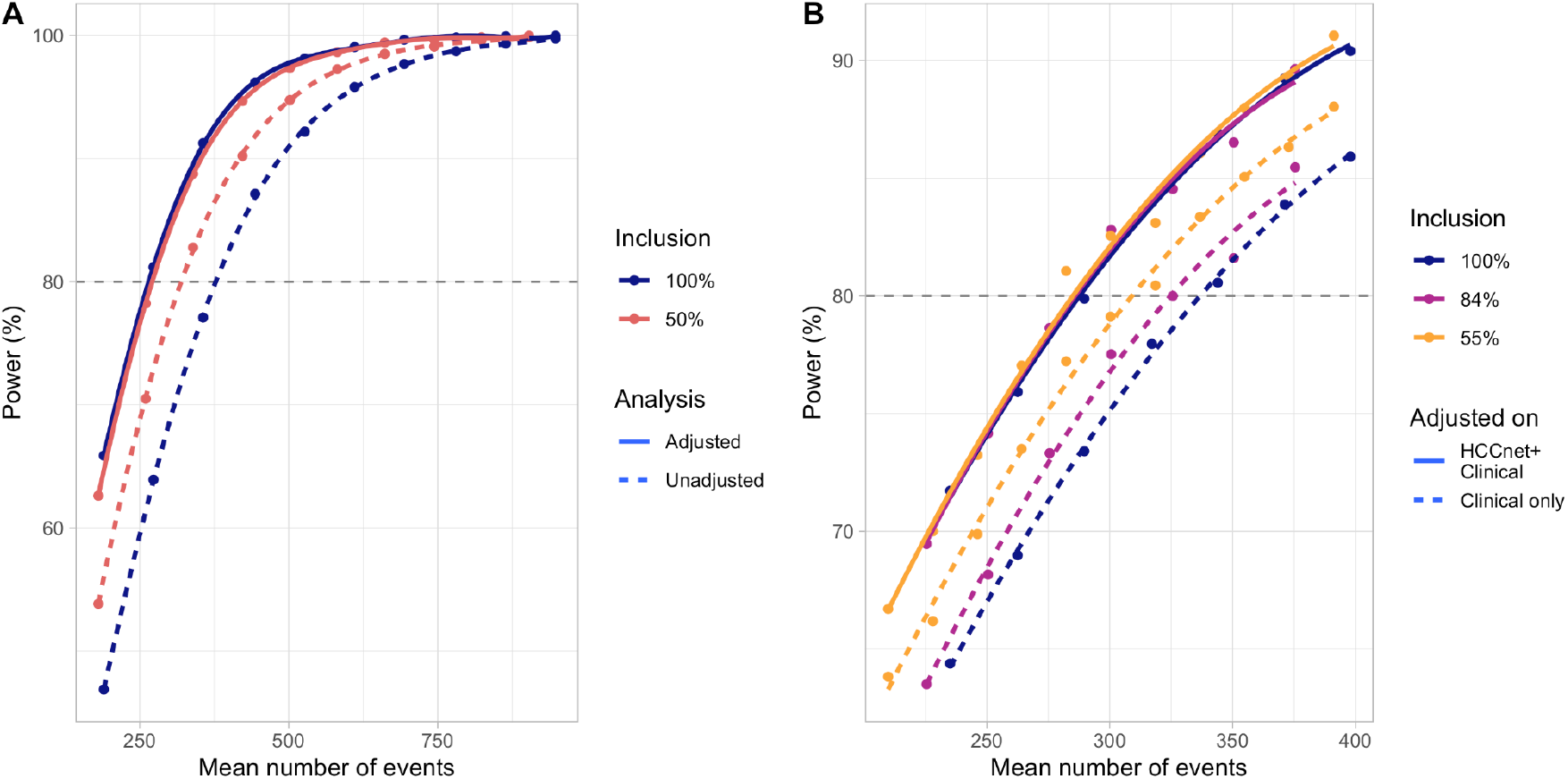
Effect of broader eligibility criteria and of covariate adjustment on statistical power. A: results of the parametric simulations where the least at risk patients are selected in the 50% inclusion scenario. B: results of the semi-synthetic simulation based on the HCC-TCGA cohort. The three levels of inclusion are based on eligibility criteria of past and ongoing trials outlined in table 1.

While the adjusted analyses with different eligibility criteria have the same statistical power, they imply a very different screened population size. Screened individuals are patients for which eligibility criteria is evaluated to test if they can be enrolled in the clinical trial. In the HCC example, the required size of the screened population is 635 for the less restrictive inclusion while it is 1690 for the most restrictive population. Therefore, the size of the screened population is divided by 2.7 when broadening eligibility criteria while attaining the same statistical power. This difference is explained by the smaller proportion of patients included as well as the smaller proportion of events with the restrictive eligibility criteria (35% at 5 years versus 50% in the entire population).

### Fit with R^2^ measures from the literature

We compare various *R*^2^measures for time-to-event endpoints to the reduction of sample size 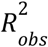 provided by covariate adjustment for the grid of parameters considered in parametric simulations (Figure S3). Most measures do not depend on the cumulative incidence of the event at the end of follow-up (Figure S3), which is not compatible with the results found for the reduction of sample size provided by covariate adjustment (Figure 2). Most measures increase only as a function of the C-index (Figure S3). The Cox-Snell 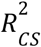 best captures the observed sample size reduction in all our simulations. The median absolute error is minimal for the 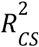, and is of 2.0% (first and third quartiles are 0.9% and 3.6% respectively). Median absolute error for other *R*^2^ measures are: 4.3% for 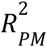 (2.0%-10.2%), 4.8% for 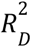 (2.2%-11.0%), 6.5% for 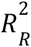 (2.5%-16.2%), 8.8% for 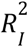 (3.8%-19.1%), 9.2% for 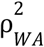 (3.7%-19.0%), 10.0% for 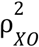 (3.5%-22.0%) and 11.2% for 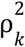 (3.9%-26.1%).

Using the Fleiss formula and the Cox-Snell 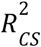 measure, we find that further adjusting on HCCnet–in addition to clinical covariates–in a resected HCC trial would decrease the sample size by 7.4%, which is an underestimation of the 16.4% reduction in sample size found with the semi-synthetic simulations.

## Discussion

The impact of covariate adjustment depends on several characteristics related to indications and clinical trials. Our simulations confirm the expected result that the power gains increase with the prognostic performance, measured by C-index, of the covariates used in covariate adjustment. Other parameters that were considered such as Weibull shape, drop-out rate or effect size do not play an important role in determining power gain. Previous work on the topic already identified that the drop-out rate and effect size do not impact the precision gains obtained with covariate adjustment[2].

Cumulative incidence at the end of the follow-up period is another major determinant of the impact of covariate adjustment. Compared to earlier work[2], we considered a finite time horizon (i.e. follow-up of 5 years) which allowed us to identify the strong dependence on cumulative incidence. Dependence on cumulative incidence is related to the dependence on the prevalence of events that occur for binary outcomes[3]. We investigated cumulative incidence for several published trials in oncology. Covariate adjustment will have limited impact for trials of new endocrine therapies for early breast cancer. For indications with low cumulative incidence, prognostic information can be more useful to perform prognostic enrichment than for covariate adjustment[40]. For aggressive cancers such as mesothelioma, metastatic breast cancer or metastatic pancreatic cancer, covariate adjustment provides notable gains in precision.

Another advantage of covariate adjustment is that it removes incentive to homogeneize the population with restrictive eligibility criteria. In the two simulation scenarios, the adjusted analysis is just as powerful whether there are strict eligibility criteria or not. However, the size of the population that needs to be screened for inclusion can be reduced substantially with the least restrictive eligibility criteria. Adequate covariate adjustment can therefore go hand in hand with broader eligibility criteria that would allow easier enrollment as well as better generalizability of trial results. This would be in line with recent calls for less restrictive eligibility criteria[11,12].

To be able to determine the sample size reduction brought by covariate adjustment, we investigated whether several *R*^2^ measures could approximate the observed sample size reduction. We found that the Cox-Snell 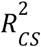 was the best approximation of our quantity of interest. The sample size with adjustment is then 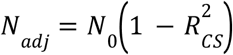 and this generalizes the Fleiss formula to a time-to-event outcome. When denoting *n* the number of patients, and *l*_0_ and *l*_1_ the log-likelihoods of a base model and a model adjusting for additional covariates, we have 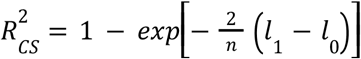 [31]. Other *R* measures we consider were developed such as they do not depend on cumulative incidence explaining why they cannot approximate the reduction of covariate adjustment provided by covariate adjustment[17,18].

Having an approximation for sample size determination is of practical importance as it can help to design clinical trials. It can also be useful in the case of a blinded sample size reestimation when there is uncertainty on the prognostic performance of adjustment covariates and where the required number of events should be reevaluated at an interim stage. Blinded sample size reestimation procedures have been proposed for a continuous outcome and could be generalized for time-to-event outcomes[41].

As noted in the draft FDA guidance, covariate adjustment changes the target of estimation, a phenomenon called non-collapsibility[8]. When adjusting for a prognostic covariate and when there is a true treatment effect, it is expected that the conditional estimand (e.g. hazard ratio) is drifted away from 1 compared to the marginal estimand but statistical uncertainty is increased. Because the amount of drift is superior to the inflation of uncertainty, statistical power resulting from covariate adjustment is increased as found in our simulations[42]. If a marginal estimand is preferred, one can consider adjusted marginal estimators that target the estimand of the unadjusted analysis while leveraging the gain in precision offered by covariate adjustment[42,43].

Our simulations study the effect of covariate adjustment on a relative measure of treatment effect, which is the hazard ratio. Absolute measures of efficacy such as restricted mean survival time or absolute risk reduction are also of interest. Estimation of those measures can also be improved by using the prognostic signal of covariates[44–46]. The extent to which our findings, for instance the dependence on cumulative incidence, generalize to this setting should be studied in further work.

Overall, we have shown that covariate adjustment reduces the sample size that is needed to reach a targeted statistical power. Reduction is particularly pronounced for indications where cumulative incidence is large. Furthermore, adequate covariate adjustment allows to maintain statistical power while relaxing eligibility criteria. New sources of prognostic covariates such as deep-learning models based on images can lead to more efficient trials.

## Data Availability

TCGA data is available by download except for predictions of HCCnet that are available upon reasonable request to the authors.

https://portal.gdc.cancer.gov/

## Supplementary material

### Supplementary Tables

**Table S1:** Description of simulation parameters used for parametric simulations of the time-to-event model.

| Quantity | Notation | Values |
| --- | --- | --- |
| Cumulative incidence | $\Lambda$ | {0.1, 0.2, 0.3, 0.4, 0.5, 0.6, 0.7, 0.8, 0.9} |
| C-index | $C$ | {0.55, 0.65, 0.75, 0.85} |
| Weibull shape | $w$ | {0.5, 1, 1.5} |
| Drop-out rate | $d$ | {0.01, 0.1} |
| Treatment hazard ratio | $hr$ | {0.4, 0.7} |

### Supplementary Figures

**Figure S1:**
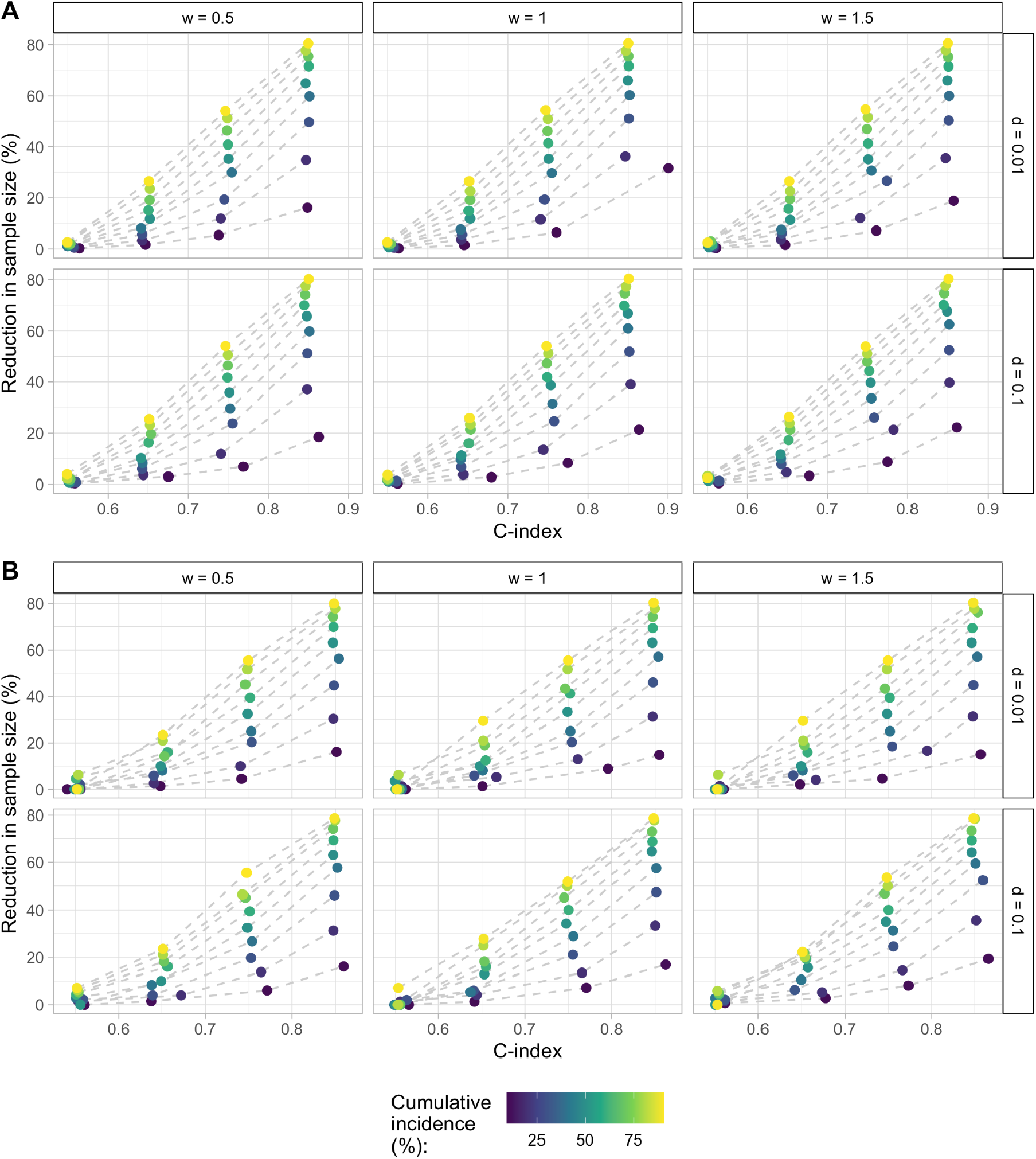
Evolution of R^2^_obs_ as a function of C-index, cumulative incidence, treatment effect, Weibull shape *w* and drop-out rate *d*. A: *hr* = 0. 7, B: *hr* = 0. 4.

**Figure S2:**
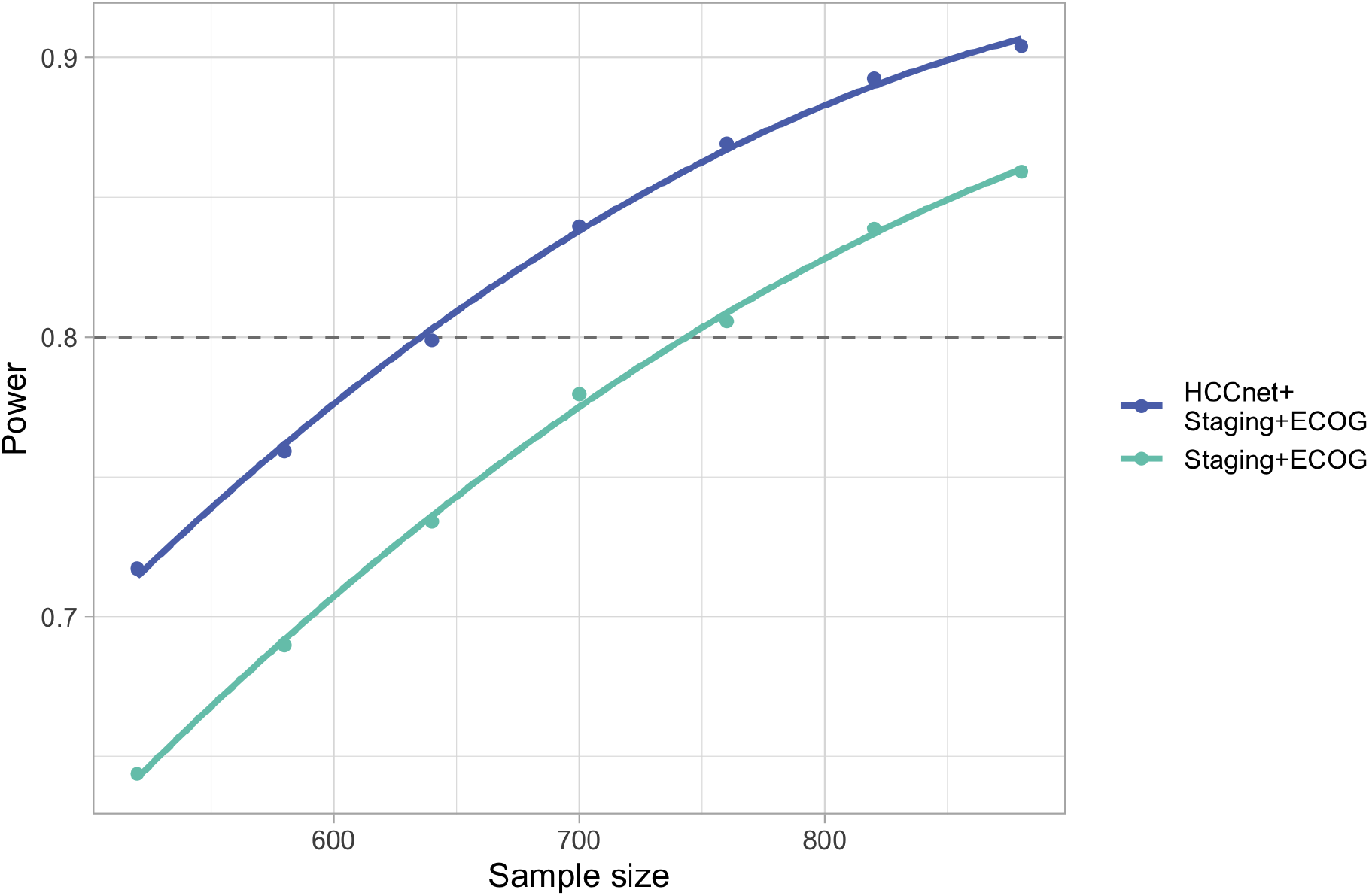
Power curves resulting from adjustment with clinical variables only (tumor staging and ECOG score) or with the additional deep learning HCCnet covariate. Covariates are sampled from the HCC patients of the TCGA dataset.

**Figure S3:**
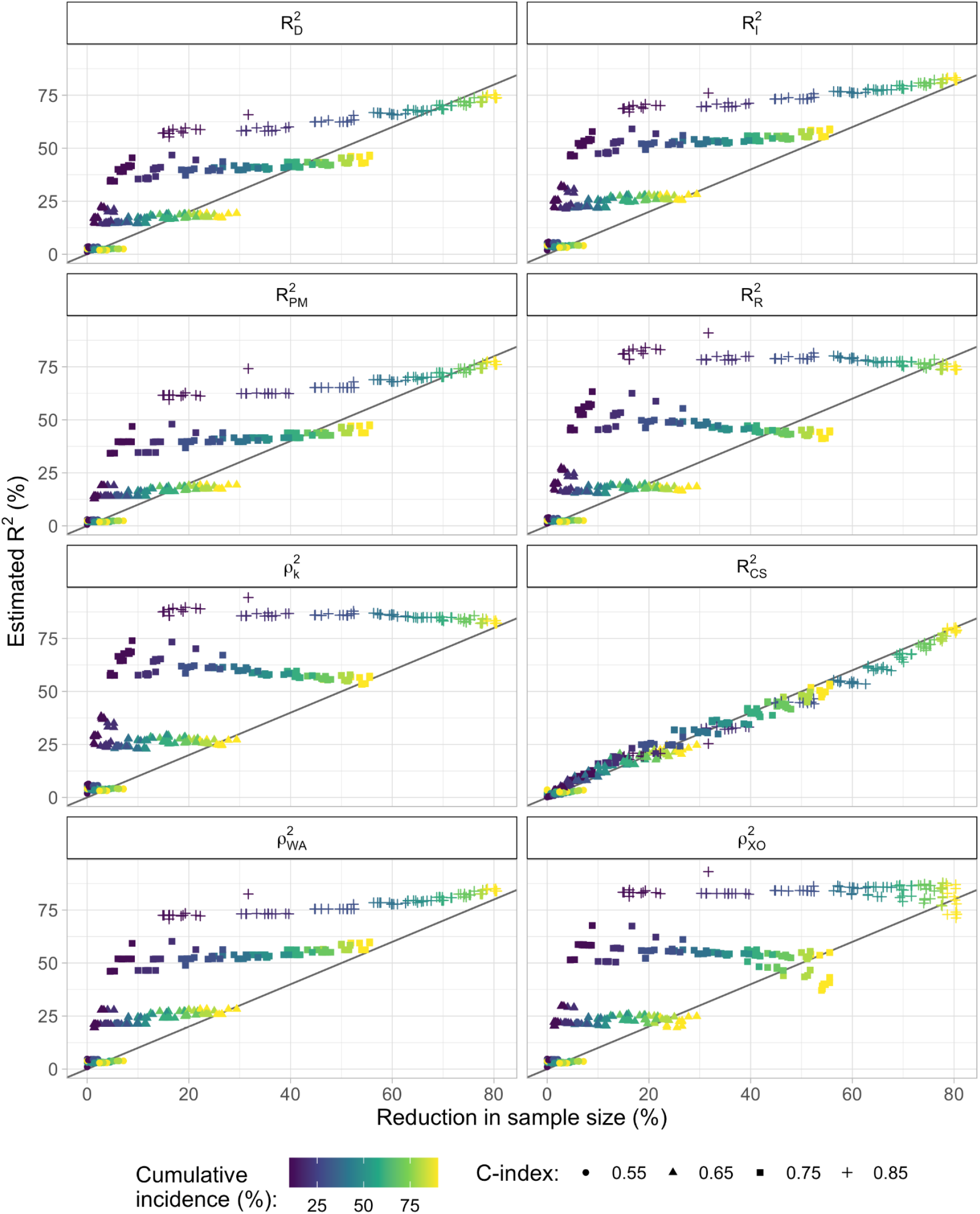
Relationships between proposed R^2^ measures and the reduction of sample size provided by covariate adjustment R^2^_obs_ over the grid of parameters described in table S1.

